# Online COVID-19 diagnosis with chest CT images: Lesion-attention deep neural networks

**DOI:** 10.1101/2020.05.11.20097907

**Authors:** Bin Liu, Xiaoxue Gao, Mengshuang He, Fengmao Lv, Guosheng Yin

**Affiliations:** Center of Statistical Research, School of Statistics, Southwestern University of Finance and Economics, Chengdu, China; Department of Statistics and Actuarial Science, The University of Hong Kong, Hong Kong, China

## Abstract

Chest computed tomography (CT) scanning is one of the most important technologies for COVID-19 diagnosis and disease monitoring, particularly for early detection of coronavirus. Recent advancements in computer vision motivate more concerted efforts in developing AI-driven diagnostic tools to accommodate the enormous demands for the COVID-19 diagnostic tests globally. To help alleviate burdens on medical systems, we develop a lesion-attention deep neural network (LA-DNN) to predict COVID-19 positive or negative with a richly annotated chest CT image dataset. Based on the textual radiological report accompanied with each CT image, we extract two types of important information for the annotations: One is the indicator of a positive or negative case of COVID-19, and the other is the description of five lesions on the CT images associated with the positive cases. The proposed data-efficient LA-DNN model focuses on the primary task of binary classification for COVID-19 diagnosis, while an auxiliary multi-label learning task is implemented simultaneously to draw the model’s attention to the five lesions associated with COVID-19. The joint task learning process makes it a highly sample-efficient deep neural network that can learn COVID-19 radiology features more effectively with limited but high-quality, rich-information samples. The experimental results show that the area under the curve (AUC) and sensitivity (recall), precision, and accuracy for COVID-19 diagnosis are 94.0%, 88.8%, 87.9%, and 88.6% respectively, which reach the clinical standards for practical use. A free online system is currently alive for fast diagnosis using CT images at the website https://www.covidct.cn/, and all codes and datasets are freely accessible at our github address.

## 1 Background

The novel coronavirus disease 2019 (COVID-19) is undergoing an unprecedented global outbreak. On March 11, 2020, COVID-19 was declared as an international public health emergency by the World Health Organization (WHO). By May 26, 2020, more than 200 countries or territories have been affected by COVID-19 with a total of more than 5.6 million confirmed cases and over 348,000 deaths. Both the numbers of confirmed cases and deaths continue climbing up quickly worldwide.

The fast increasing numbers of COVID-19 cases and deaths have caused overburdens on many local medical systems across the world. Currently, the reverse-transcription polymerase chain reaction (RT-PCR) test is the standard method for detecting the coronavirus in COVID-19 patients. However, the laboratory RT-PCR test usually takes a rather long time (in days) to deliver the final result. To shorten the time of diagnosis, the real time RT-PCR test is recommended which can deliver a reliable diagnosis result much faster (in 6–8 hours). Such tests need a collection of a sample with a swab that goes deep in a person’s nose or throat, i.e., parts of the body where the coronavirus gathers. However, if the swabbed areas do not have coronavirus accumulated, the tests may fail to identify a COVID-19 patient correctly. Many countries are experiencing a backlog of test results due to a lack of diagnostic kits at their medical facilities, and the test results may even take longer time than anticipated due to the increasing demands for testing globally. Not only are these tests insufficient to meet the urgent and vast demands in many countries (particularly those with poor medical infrastructures), but they are also inefficient as the time lag of test reporting may cause treatment delay, especially for patients with critical conditions. Moreover, the sensitivity of the current RT-PCR testing kits is not high; that is, a large number of COVID-19 patients cannot be identified accurately after their first tests due to false negatives. As a result, it usually requires several tests to make a final confirmation. Hence, patients may not receive appropriate treatment and necessary quarantine during the RT-PCR testing period. On the other hand, chest CT scan is another critical tool for COVID-19 diagnosis and disease monitoring, particularly for early detection when the symptoms are yet onset. After entering the body, coronavirus often first attacks the lung and thus certain lesions would manifest before a swab can collect virus in the nose or throat. According to many existing studies [1, 3], CT scanning serves as a important and necessary supplement for the RT-PCR test and sometimes can even outperform the laboratory test for COVID-19 diagnosis. In contrast to the RT-PCR test, the chest CT scans are common and the corresponding diagnostic results can be obtained in a much faster way.

To improve the efficiency of the CT-based diagnosis, automatic diagnostic systems have been developed with AI technologies by reading patients’ chest CT images as inputs and then output the diagnosis results [6, 9, 11, 13]. These AI-driven methods have demonstrated very promising performances on COVID-19 prediction. However, most of the existing work do not share the training data publicly, while He et al. [6] constructed the first openly accessible COVID-19 chest CT dataset by extracting the CT images from over 760 preprints in *medRxiv* and *bioRxiv*. This publicly available dataset contains 746 samples, among which 345 of them are COVID-19 positive and the rest 401 are negative^1^. We keep expanding the dataset by continuously collecting new samples appeared in the latest publications on COVID-19.

Figure 1 shows the CT images in the dataset annotated with professional textual analysis, accompanied with the radiological reports in the right side. The text reports usually narrate the results on whether the patients are COVID-19 positive or not. In addition to the flag of COVID-19, the text also contains information on descriptions of lesions of COVID-19 patients. Based on our comprehensive statistical analysis over the entire text annotations, there are five different lesions associated with COVID-19, including the Ground glass opacity (GGO), Consolidation (Csld), Crazy paving appearance (CrPa), Air bronchograms (AirBr), and Interlobular septal thickening (InSepThi). Figure 1 shows that each of the confirmed COVID-19 cases is attached with one up to five lesion labels. Our experiments corroborate that the auxiliary information on these lesions is extremely valuable for COVID-19 diagnosis and can greatly improve the diagnostic accuracy. However, the pioneering work [6] only focused on the COVID-19 diagnosis by conducting a binary classification task on predicting the flag of COVID-19, but ignored the significant amount of information on the common lesions which are distinctive from other types of pneumonia.

**Figure 1:**
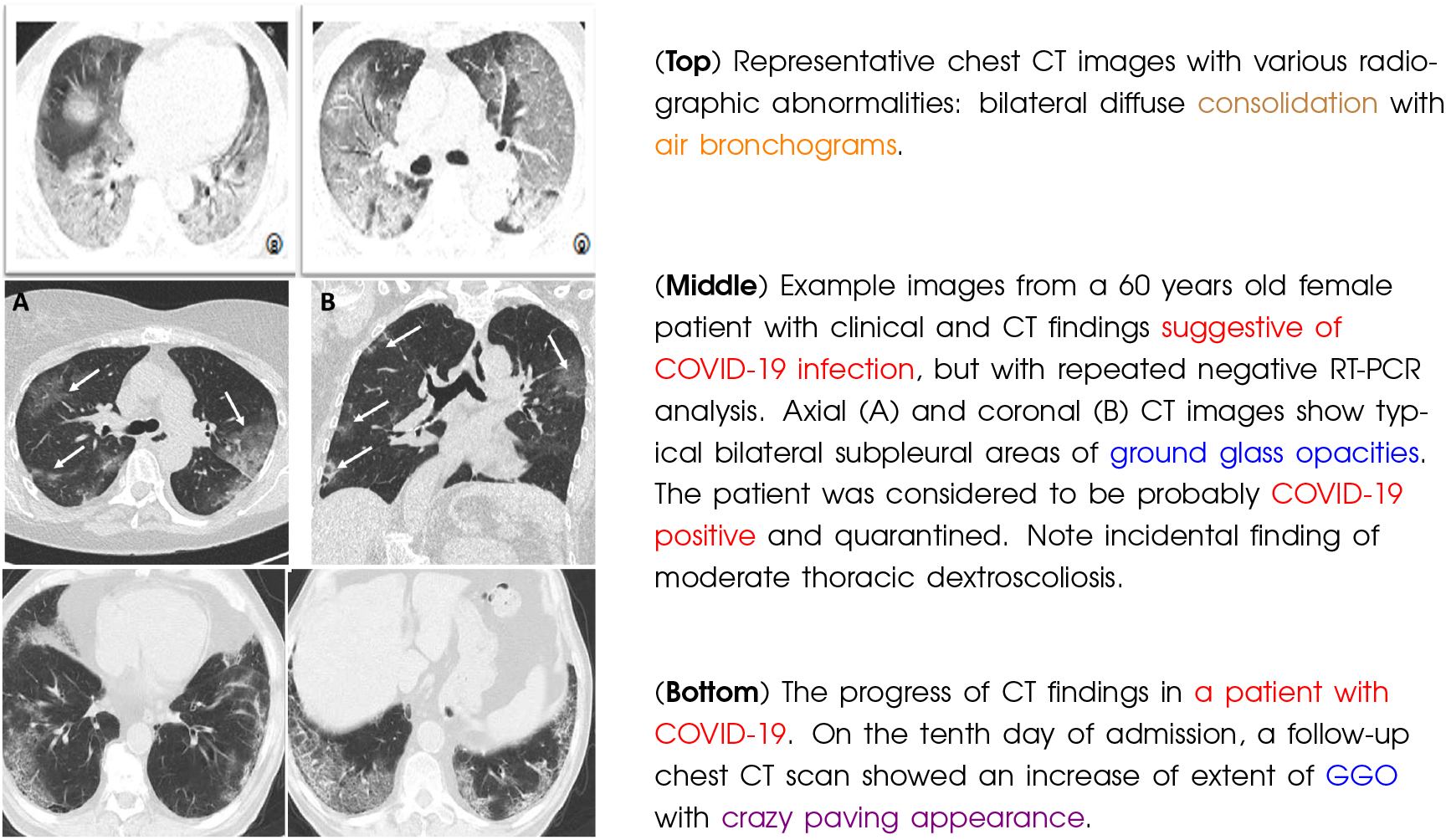
Illustration of the multi-label chest CT images collected from online papers. All of them have been confirmed as COVID-19 positive. We highlight the six labels with different color: COVID-19 in red, Ground glass opacities (GGO) in blue, Consolidation (Csld) in brown, Crazy paving appearance (CrPa) in violet, Air bronchograms (AirBr) in orange, and Interlobular septal thickening (InSepThi) in magenta. Taking the CT scans of the second patient (Middle) as an example, these chest CT images have been annotated with three labels: COVID-19, GGO, and InSepThi.

We develop a highly accurate COVID-19 diagnosis system based on the chest CT images as well as the corresponding rich annotations on the five lesions. Our model adopts a double-task learning process which contains a primary binary classification task on the flag of COVID-19 and an auxiliary multi-label attention learning task on the five lesions. Both tasks are trained synchronously, while it shows that the auxiliary task promotes the primary task to focus its attention on the lesion areas and, as a result, the diagnostic accuracy of COVID-19 is improved above the level of the state-of-the-art method. Due to the incorporation of the attention mechanism on the five lesions, we refer to our new model as the lesion-attention deep neural network (LA-DNN).

Experimental results demonstrate that our LA-DNN model can achieve great improvements by using the textual information. The area under the curve (AUC), recall (sensitivity), F1 score, and accuracy for predicting the diagnosis of COVID-19 are 94.0%, 88.8%, 87.9%, and 88.6%, respectively. These results improve drastically beyond the existing work and reach the clinical standards for COVID-19 diagnosis [3, 8]. Therefore, our system can be deployed for practical use to alleviate the enormous burdens of COVID-19 diagnostic tests [3]. The annotated lesion label file and the implementation codes in Python can all be freely accessed at https://github.com/xiaoxuegao499/LA-DNN-for-COVID-19-diagnosis. An online system has been developed and is openly available for fast COVID-19 diagnosis using chest CT images at the website https://www.covidct.cn/. Our online system also welcomes medical staff to upload new patients’ CT image data to validate the diagnostic results, as well as keeping the system routinely updated with the data shared publicly.

## 2 Methodology

### 2.1 Motivation and model

There has been an increasing amount of work on developing an AI-based COVID-19 diagnostic system using patients’ chest CT scans [11]. Unfortunately, most of the data used in the deep learning models are not publicly available, which makes the existing models and results difficult to verify and reproduce. He et al. [6] published the first open-access COVID-19 chest CT scans dataset based on the CT images appeared in online preprints of research work on COVID-19. In a supervised learning process, classification based on deep learning models typically requires a relatively large number of annotated samples to train the model for accurate prediction. However, the current publicly available dataset [6] only contains 746 public chest CT images. The shortage of labeled samples and the urgency for the development of automated COVID-19 diagnostic tools motivate us to derive a sample-efficient deep model that can integrate all sources of information for optimal decision making.

Through careful studies on the preprints associated with the patients’ chest CT scans, we can extract valuable textual annotations of these CT scans. One is the flag of COVID-19, and the other is the radiological reports on five potential lesions in the lung. In the pioneering work by He et al. [6], they trained a binary classification model based on the COVID-19 flag only while ignoring all the lesion information which requires further annotations. To improve the performance of diagnosis, we propose to integrate the information on the lesion descriptions into the classification of the flag of COVID-19.

Our goal is to make accurate classification of the COVID-19 positive or negative directly using the chest CT images. However, different from the work of He et al. [6] which focused on making a complex knowledge transfer, we aim to fully exploiting the richly annotated textual information in the data. After annotating the five-category lesions on the COVID-19 positive images, we propose an auxiliary multi-label learning model based on the summarized five different lesion labels, in addition to the primary objective of the binary classification for COVID-19. The auxiliary task applies multi-label learning over the five lesions annotated based on the corresponding radiological reports: Ground glass opacities (GGO), Consolidation (Csld), Crazy paving appearance (CrPa), Air bronchograms (AirBr), and Interlobular septal thickening (InSepThi), as shown in Figure 2. The primary and auxiliary tasks are trained synchronously in our LA-DNN model, so that the unknown parameters can be learned much more effectively than those by only training the primary task for binary classification. The auxiliary multi-label learning task promotes the fine-grained information on the radiology-revealed lesions to be integrated into the primary task, which makes the primary task pay more attention to the lesion areas rather than other uninteresting areas when making a final decision. This lesion-attention mechanism drastically improves the diagnostic accuracy up to the level of clinical standards by medical experts.

**Figure 2:**
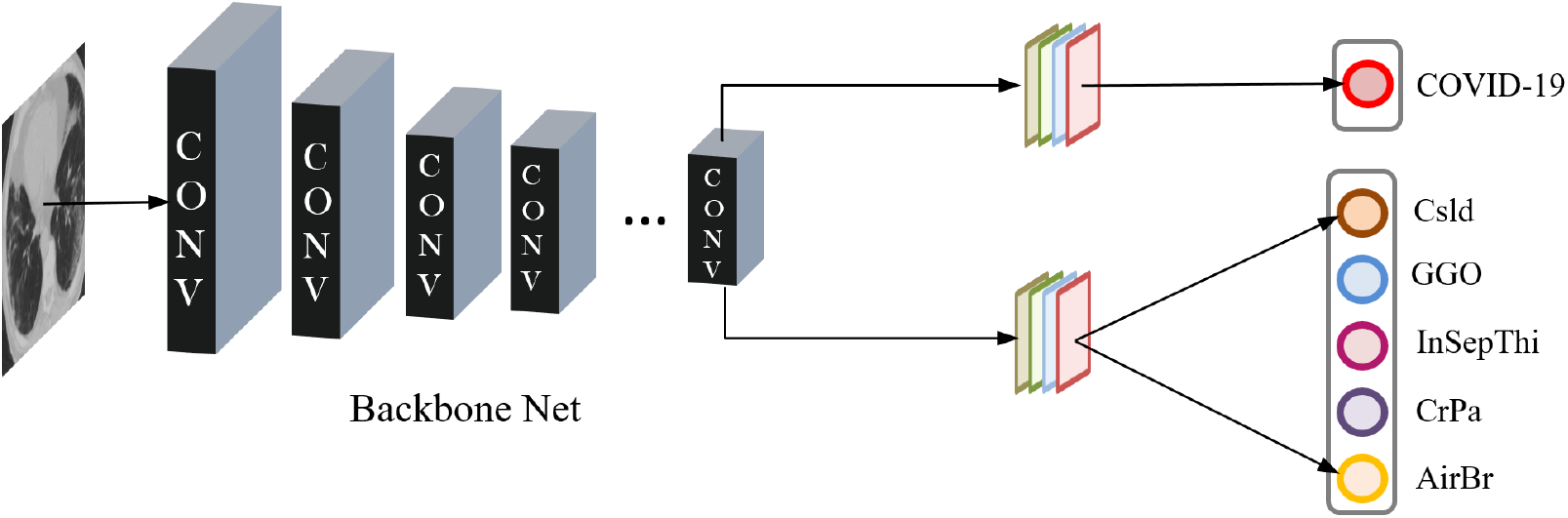
The architecture of the proposed lesion-attention deep neural networks with a primary task of binary classification and an auxiliary task of multi-label learning of five lesions.

### 2.2 Implementation of LA-DNN

Figure 2 shows the architecture of the proposed LA-DNN model. Using the ImageNet, we first pre-train the backbone networks, and then via the idea of transfer learning, a pre-trained backbone network takes the patient’s chest CT images as inputs. Seven well-known deep neural networks are explored one at a time to be used as the backbone networks in the experiments, including VGG-16 [10], ResNet-18 [5], ResNet-50 [5], DenseNet-121 [7], DenseNet-169 [7], EfficientNet-b0 [12], and EfficientNet-b1 [12]. The output of the last layer of the backbone networks is carried forward to two branches. One is used to predict whether a patient is COVID-19 positive or not, which is the primary task. Simultaneously, the other branch aims to make a multi-label prediction on the five lesions. The prediction errors from both the primary binary task and the auxiliary multi-label task are fed back to fine-tune the backbone network.

### 2.3 COVID-19 chest CT scans

We train the proposed LA-DNN model on the dataset published by He et al. [6] in conjunction with our newly collected dataset. This original dataset contains 345 samples of COVID-19 positive and 401 COVID-19 negative CT scans, which are collected from 760 research preprints related to COVID-19 from *medRxiv* and *bioRxiv*, posted from January 19th to March 25th 2020. We keep expanding the dataset of chest CT images by further collecting 219 new positive samples from 57 COVID-19 confirmed patients and 259 new negative samples from 164 non-COVID-19 patients from the newly appeared publications up to May 21th. We denote the original dataset with 746 samples as 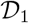, and denote the dataset with newly collected 483 samples as 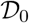. The combined dataset 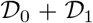 contains a total of 564 (269 patients) positive samples and 660 (339 patients) negative samples. Table 1 illustrates the details of data splitting.

**Table 1:** Stratified by COVID-19 positive or negative, the number of samples (CT images) and the number of patients (in parentheses) in each dataset based on the patient ratios of data splitting 60%, 15%, 25% for training, validation, and testing sets.

|  | COVID-19 |  | COVID-19 + | COVID-19 - |
| --- | --- | --- | --- | --- |
| Datasets | + | - | Training/Validation/Testing | Training/Validation/Testing |
| $\mathcal{D}_0$ | 345(212) | 401(175) | 187(126)/60(32)/98(54) | 238(109)/58(24)/105(42) |
| $\mathcal{D}_1$ | 219(57) | 259(164) | - | - |
| $\mathcal{D}_0 + \mathcal{D}_1$ | 564(269) | 660(339) | 340(169)/81(34)/143(66) | 398(215)/97(44)/165(80) |

The negative samples of the combined dataset include the CT scans of normal individuals as well as patients with other types of diseases, e.g., lung cancer or other types of pneumonia. The COVID-19 positive CT images are further annotated with the corresponding radiological reports, which are the textual clinical conclusions for the patients. The radiological information is proven to be extremely valuable for the COVID-19 diagnosis. From the textual annotations, we extract two types of important information:

- The first type of information is on whether patients are diagnosed positive or negative for COVID-19, which corresponds to the binary classification labels;
- The second significant but often ignored information is a common description of the five lesions associated with COVID-19. Each CT image of a confirmed COVID-19 patient has been identified with one up to five lesions in the lung as shown in Figure 1.

Figure 3 shows the visualization of the positive and negative samples in our dataset. Specifically, Figure 3 (a) and (b) exhibit the distribution of the five lesion labels and their concordance matrix (i.e., the number of times two lesions appeared in the same sample). From the histogram of the numbers of labels for all positive samples, we find that most of the positive samples have either one or two lesion labels. The paired-label concordance matrix of the 212 COVID-19 positive samples which have two lesion labels demonstrates that GGO and consolidation often appear together in the CT images, as shown in Figure 3 (b). Moreover, GGO are the lesion that has the most frequent interactions with all the other four lesions. Figure 3 (c) illustrates the composition of the COVID-19 negative samples. The negative sample set contains lung cancer, lung nodules (LNodules), pulmonary viral pneumonia (PVP) which is non-COVID-19, and normal samples. The lung cancer covers many types of common cancers including lung adenocarcinomas (LungAden), lung squamous cell carcinoma (LungSCC), and others.

**Figure 3:**
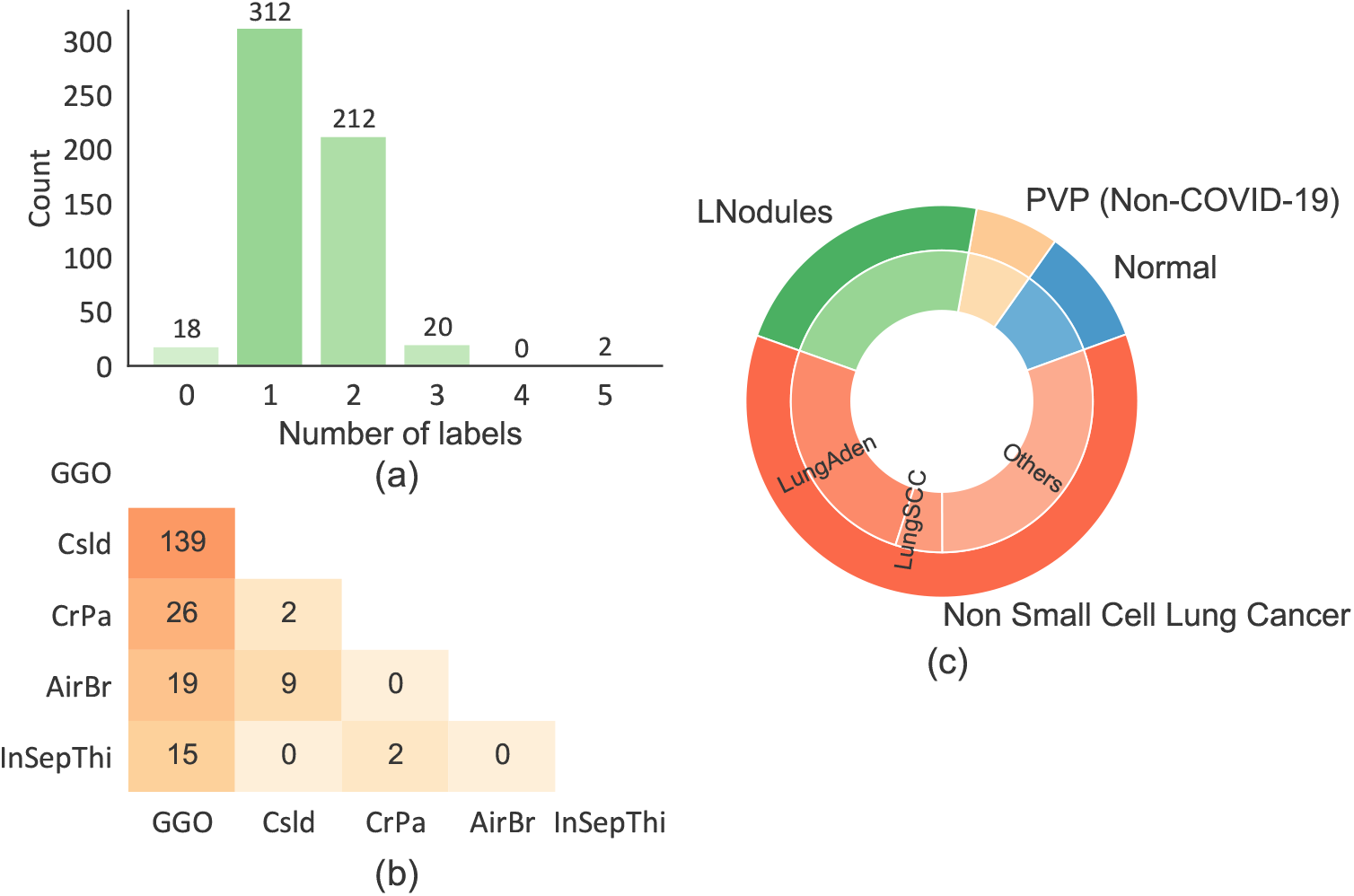
Descriptions of COVID-19 positive and negative samples: (a) Histogram of the numbers of lesions in the COVID-19 positive set; (b) Lesion concordance matrix; and (c) Composition of the COVID-19 negative samples.

## 3 Results

### 3.1 Overall performance

In our experiments, we split both the original dataset 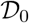 and the combined dataset 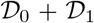 respectively into a training set, a validation set, and a testing set by patients’ IDs with the same ratios of 60%, 15%, 25% as those in [6]. One patient may possibly has multiple CT images in the dataset, while all CT images belonging to the same patient would be allocated together to the same set. We need to ensure that the COVID-19 patients in the training, validation, and testing sets cover the one to five lesion labels based on the new annotations using the radiological reports.

Given the limited training samples, we first take a classical neural network that has been well pre-trained on a large dataset ImageNet [4] as a feature extraction function, and then fine-tune the weights with the COVID-19 chest CT dataset. We select seven popular architectures as the backbone networks, including VGG-16 [10], ResNet-18 [5], ResNet-50 [5], DenseNet-121 [7], DenseNet-169 [7], EfficientNet-b0 [12], and EfficientNet-b1 [12].

We take the pioneering work of He et al. [6] as the baseline for comparison. We train the baseline and the proposed LA-DNN model on the original dataset 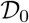 as well as the combined dataset 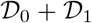 with the same data splitting strategies. Tables 2 summarizes the overall performances of the baseline and our LA-DNN model with different backbone networks. Clearly, the proposed LA-DNN model significantly improves the prediction of COVID-19 positive or negative on all of the four metrics with both datasets. Experimental results demonstrate that the auxiliary task learning process by using the five lesions from the textual information can greatly improve the primary task for binary classification of COVID-19. Moreover, the additional dataset 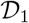 can further improve the model’s prediction accuracy. Among all the methods considered, the best performance is delivered by the LA-DNN using the VGG-16 as the backbone net with the combined dataset. The corresponding AUC, recall (sensitivity), precision, and accuracy for predicting the diagnosis of COVID-19 patients are 94.0%, 88.8%, 87.9%, and 88.6%, which reach the clinical standards for COVID-19 diagnosis in practice [3, 8].

**Table 2:** Comparisons of sensitivity, AUC, F1 score, and accuracy (%) between the baseline model and our LA-DNN model on the original dataset 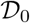 and the combined dataset 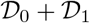.

| Metrics | Backbone Net | $D_0$ | | $D_0 + D_1$ | |
| --- | --- | --- | --- | --- | --- |
|  |  | Baseline | LA-DNN | Baseline | LA-DNN |
| Sensitivity | VGG-16 | 75.5 | 84.7 | 85.3 | **88.8** |
|  | ResNet-18 | 71.4 | 80.6 | 75.5 | 82.5 |
|  | ResNet-50 | 67.3 | 78.6 | 82.5 | 87.4 |
|  | DenseNet-121 | 77.6 | 79.6 | 86.0 | 86.0 |
|  | DenseNet-169 | 77.6 | 85.7 | 85.3 | 87.4 |
|  | EfficientNet-b0 | 68.4 | **87.8** | 86.7 | 88.1 |
|  | EfficientNet-b1 | 70.4 | 77.6 | 76.2 | 86.0 |
| AUC | VGG-16 | 81.3 | 89.8 | 92.7 | 94.0 |
|  | ResNet-18 | 83.0 | 88.2 | 93.1 | 93.1 |
|  | ResNet-50 | 87.6 | 90.5 | 91.6 | 93.8 |
|  | DenseNet-121 | 86.0 | 88.7 | 92.8 | 93.2 |
|  | DenseNet-169 | 88.2 | **91.2** | 93.1 | 93.3 |
|  | EfficientNet-b0 | 87.7 | 90.0 | 92.0 | 92.4 |
|  | EfficientNet-b1 | 84.1 | 87.6 | 91.6 | **94.7** |
| F1 score | VGG-16 | 74.8 | 83.0 | 84.1 | **87.9** |
|  | ResNet-18 | 74.0 | 79.8 | 80.6 | 84.6 |
|  | ResNet-50 | 75.9 | 83.2 | 82.8 | 86.8 |
|  | DenseNet-121 | 80.0 | 81.2 | 84.2 | 85.4 |
|  | DenseNet-169 | 81.3 | **84.8** | 86.2 | 86.8 |
|  | EfficientNet-b0 | 74.9 | 83.9 | 85.2 | 86.6 |
|  | EfficientNet-b1 | 73.4 | 80.0 | 79.8 | **87.9** |
| Accuracy | VGG-16 | 75.4 | 83.2 | 85.1 | 88.6 |
|  | ResNet-18 | 75.9 | 80.2 | 83.1 | 86.0 |
|  | ResNet-50 | 79.3 | 84.7 | 84.1 | 87.7 |
|  | DenseNet-121 | 81.3 | 82.3 | 85.1 | 86.4 |
|  | DenseNet-169 | 82.8 | **85.2** | 87.3 | 87.7 |
|  | EfficientNet-b0 | 77.8 | 83.7 | 86.0 | 87.3 |
|  | EfficientNet-b1 | 75.4 | 81.2 | 82.1 | **89.0** |

### 3.2 Curves for assessment

To further assess the proposed LA-DNN model, a receiver operating characteristic (ROC) curve and a precision-recall curve are exploited to evaluate performances under different threshold values when interpreting probabilistic predictions. Figure 4 demonstrates the results on the dataset 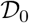. Figure 4 (a) exhibits the ROC curves of the baseline and our LA-DNN model by selecting the backbone network as VGG-16 [10] and DenseNet-169 [7] respectively. The results show that our model can predict the COVID-19 based on patients’ chest CT scans with an area under the ROC curve (AUC) of 0.912 (when choosing DenseNet-169 as the backbone net) and 0.898 (when choosing VGG-16 as the backbone net), which demonstrate significant improvements from the corresponding AUC values of 0.882 and 0.813 from the baseline. Figure 4 (b) shows the precision-recall curves of the baseline and our LA-DNN model when choosing the VGG-16 [10] and DenseNet-169 [7] as the backbone networks respectively. The precision-recall curve plots the precision and the recall under different threshold values. The ideal model with a perfect prediction corresponds to the point with the coordinates of (1,1). As shown in Figure 4 (b), the curves of our LA-DNN models using the backbone nets VGG-16 and DenseNet-169 bend towards the point (1,1) much closer than those of the baseline.

**Figure 4:**
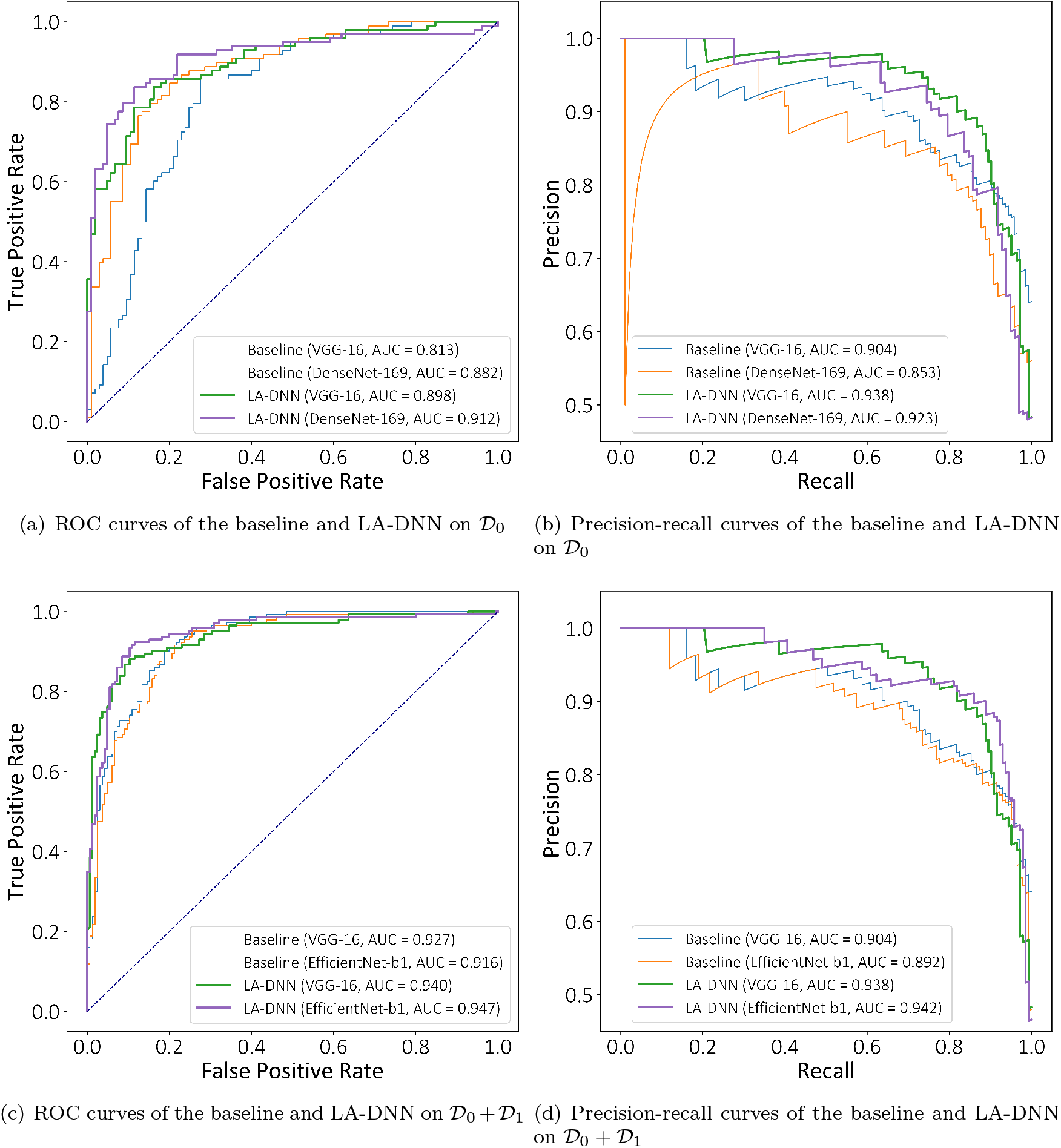
Performances of our proposed LA-DNN model for COVID-19 diagnosis in comparison with the baseline on 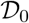 and 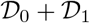

Figure 4 (c) and (d) are the results on the dataset 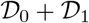. The best AUC of the ROC curve is 0.947 when choosing EfficientNet-b1 as the backbone net as shown in Figure 4 (c). Figure 4 (d) shows that the precision-recall curves of LA-DNN are much closer to (1, 1) than the baseline.

## 4 Analysis

### 4.1 Lesion attention map

To gain more insights into our LA-DNN model, we can visualize the lesion attention map (i.e., the class activation map) concerning the five lesion labels with a convolutional neural network (CNN) visualization tool, Grad-CAM++ [2]. Grad-CAM++ can localize the lesions in a CT image even if there are multiple occurrences of one lesion. Subsequently, Grad-CAM++ renders a class activation map, which illustrates the importance of each pixel in a feature map towards the final classification result. The attention heat-map exhibits the pixel-wise weighting of the gradients back-propagated from the output with respect to a particular spatial position in the final convolutional feature map of the CNN. In other words, the class activation map is a saliency map indicating which areas the model has paid attention to.

Based on our selected 10 chest CT image samples, Figure 5 shows the class-specific attention maps for the baseline and our LA-DNN model with both choosing DenseNet-169 as the backbone network. The first column represents the original COVID-19 CT images, and the lesion areas of these images are bounded with green boxes. Columns (b) and (c) in Figure 5 show the results of the baseline. In particular, column (b) is the class-specific attention map learned by the baseline. In column (c), the class attention map of the baseline is superimposed on the original images to show the activated areas. The color of the maps from deep red to dark blue corresponds to the values of pixels’ class-specific saliency from large to small. Columns (d) and (e) exhibit the corresponding results of the proposed LA-DNN model. The class-specific saliency map is a visual explanation of the lesions of COVID-19 CT scans that are predicted by the network. By comparing the results between the baseline and our model, we observe that the proposed LA-DNN model can capture almost all the salient areas for the COVID-19 prediction.

**Figure 5:**
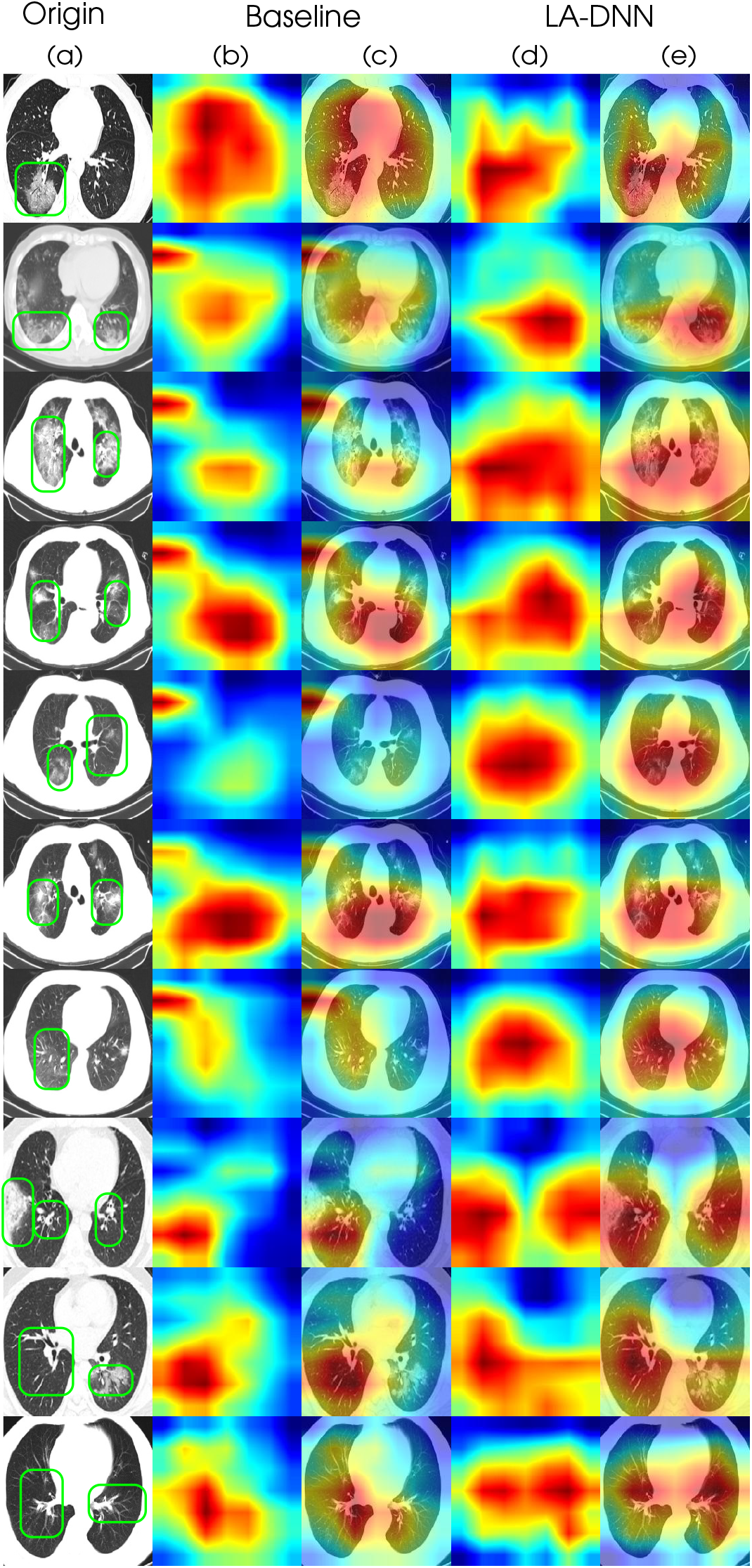
Grad-CAM++ visualization for the baseline and our LA-DNN model with the backbone net of DenseNet-169. Column (a) represents the original CT scans; Columns (b) and (c) are the class activation maps of the baseline [6]; Columns (d) and (e) are the class activation maps of our LA-DNN model. The color from deep red to dark blue corresponds to the activation values from large to small.

### 4.2 Visualization of the primary vs. auxiliary tasks

During the testing, the primary task of our LA-DNN model outputs a binary label on COVID-19 diagnosis, and meanwhile the auxiliary task outputs a five-dimensional vector to predict the labels of five lesions. Figure 6 shows the numeric components of these five-dimensional vectors paired with the COVID-19 classification labels. The paired plot creates a matrix for the five lesions of GGO, Csld, CrPa, AirBr and InSepThi. The diagonal figures exhibit the distributions of values of each lesion from the auxiliary task in distinguishing COVID-19 positive or negative. The offdiagonal axes display the distribution of each paired lesions over the two categories: COVID-19 or non-COVID-19. Not only are the GGO and Csld lesions common in COVID-19 but they also frequently appear in non-COVID-19 cases (i.e., other types of pneumonia). As a result, both lesions are less powerful in helping to triage COVID-19 or non-COVID-19. In summary, the paired plots in Figure 6 corroborate that the three lesions of CrPa, AirBr, and InSepThi are more important factors than GGO and Csld in distinguishing COVID-19 from non-COVID-19.

**Figure 6:**
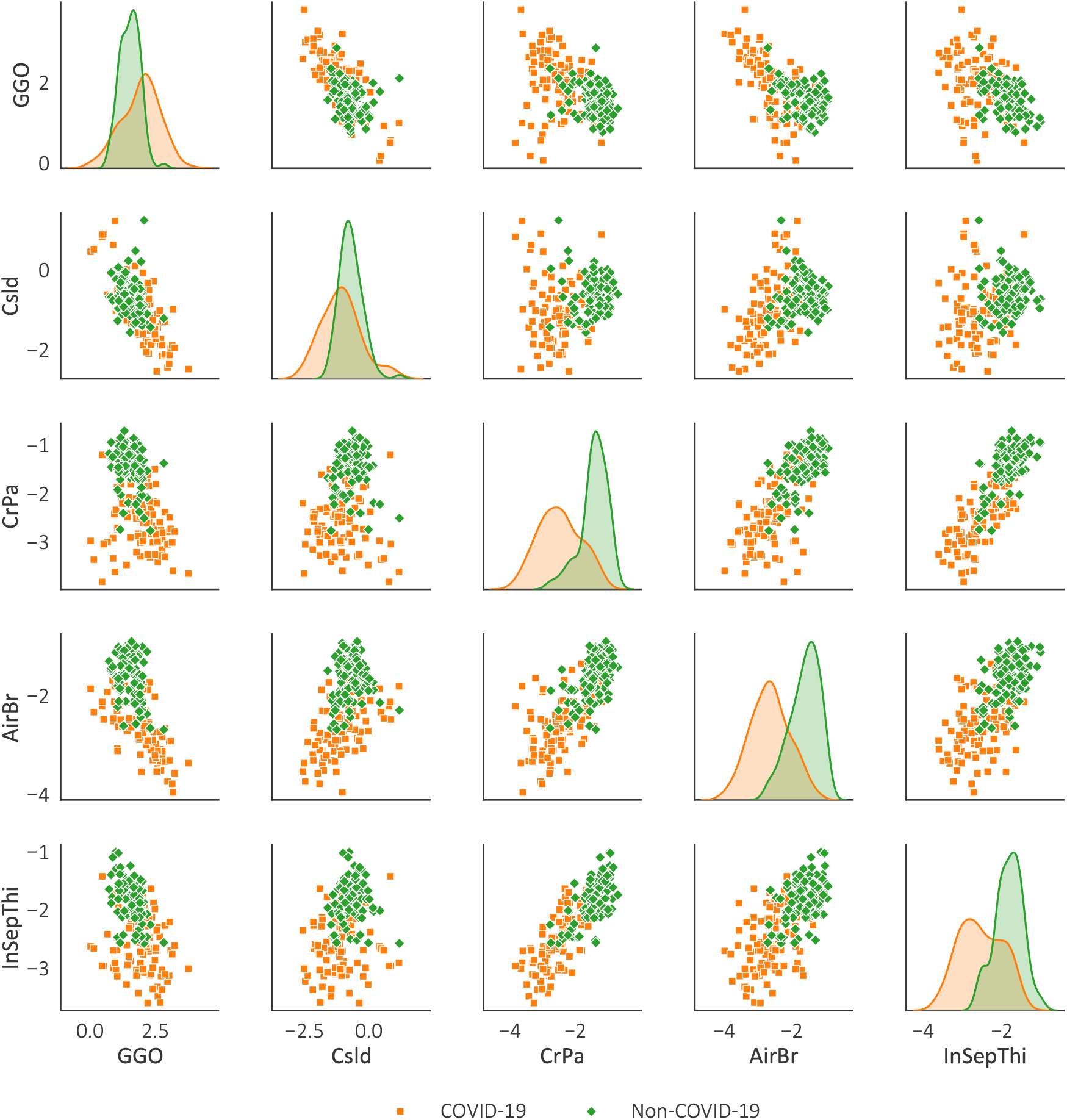
Plots of the pairwise relationships among the five lesions on making the final binary classification of COVID-19 or non-COVID-19.

## 5 Conclusion

To accommodate the urgent and enormous demands for COVID-19 testing, we develop a multi-lesion attention deep neural network for automating the COVID-19 diagnosis using richly annotated chest CT image data. The samples of our dataset are collected from over 760 preprinted papers as well as from newly added images. To overcome the limitation of the sample size, we extract two types of supervised information from the radiological text: One is the flag of COVID-19, and the other is the multiple labels for the five lesions of COVID-19. The rich annotations allow us to propose a sample-efficient deep neural network to learn valuable features with a limited number of samples. The proposed highly data-driven deep model contains a primary task on the binary classification for COVID-19 and an auxiliary multi-label attention task which forces the model to pay close attention to the five lesions of COVID-19 during the training process. The experimental results demonstrate that the proposed LA-DNN model is capable of achieving the current clinical standards for diagnostic testing and thus our system should be broadly deployed for practical use. Currently, an online version of the AI-driven COVID-19 diagnostic system is set up for validation and continual collection of the data. All our codes and annotated data are publicly available to help other researchers to further develop more accurate systems to defeat the COVID-19.

## Data Availability

The data and computer codes used in this study are publicly available.

https://github.com/UCSD-AI4H/COVID-CT

https://github.com/xiaoxuegao499/LA-DNN-for-COVID-19-diagnosis

## Footnotes

1 The original dataset [6] has 349/397 positive/negative samples, while the authors assigned 4 negative samples with positive labels by mistake.

